# Multi-omics analysis of the molecular response to glucocorticoids - insights into shared genetic risk from psychiatric to medical disorders

**DOI:** 10.1101/2023.12.05.23299430

**Authors:** Janine Knauer-Arloth, Anastasiia Hryhorzhevska, Elisabeth B. Binder

**Affiliations:** Department Genes and Environment, Max Planck Institute of Psychiatry, 80804 Munich, Germany; Institute of Computational Biology, Helmholtz Munich, 85764 Neuherberg, Germany; Department of Psychiatry and Behavioral Sciences, Emory University School of Medicine, Atlanta GA 30322, USA

**Author notes:** Corresponding authors: J. Knauer-Arloth & E. B. Binder.

**Keywords:** DNA methylation, Gene expression, Genetic variation, Stress response, Multi-omics network analysis, Immune response, Cell type-specificity

## Abstract

**Background:** Glucocorticoids play a crucial role as mediators of negative health effects associated with chronic stress, including increased risk for psychiatric disorders as well as cardiovascular and metabolic diseases. This study investigates the impact of genetic variants and glucocorticoid receptor (GR)-activation on gene expression and DNA methylation in peripheral blood and the relationship of these variants with disease risk.

**Methods:** We conducted a comprehensive molecular quantitative trait locus (QTL) analysis, mapping GR-methylation (me)QTLs, GR-expression (e)QTLs, and GR-expression quantitative trait methylation (eQTM) in a cohort of 199 individuals, with DNA methylation and RNA expression data collected before and after GR-activation with dexamethasone. A multi-level network analysis was employed to map the complex relationships between the transcriptome, epigenome, and genetic variation.

**Results:** We identified 3,772 GR-meQTL CpGs corresponding to 104,828 local GR-meQTLs. eQTM and eQTL analyses revealed distinct genetic influences on RNA expression and DNA methylation. Multi-level network analysis uncovered GR-network trio QTLs, characterized by SNP-CpG-transcript combinations where meQTLs act as both eQTLs and eQTMs. These trios’ genes demonstrated enrichment in immune response and cell activation pathways and showed a significant overlap with transcripts altered by GR-activation in the mouse brain. GR-trio variants were enriched in GWAS for bipolar disorder, schizophrenia, autoimmune and cardiovascular diseases, along with associated traits, cytokines levels, and BMI.

**Conclusions:** Genetic variants modulating the molecular effects of glucocorticoids are associated with psychiatric as well as medical diseases. Our findings support stress as a shared risk factor for transdiagnostic negative health outcomes and may lead to innovative interventions targeting shared underlying molecular mechanisms.

## Introduction

Glucocorticoids (GC) are the main effectors of the hypothalamus-pituitary-adrenal (HPA) axis and play a crucial role in mediating the acute stress response. They are also implicated in the negative health effects of chronic stress, which include an increazed risk for a variety of psychiatric disorders, cardiovascular diseases, diabetes, and cancer (1). Studies have shown that chronic exposure to GCs can increase morbidity and even reduce life span (2,3). In addition, synthetic GCs are effective in the treatment of different medical disorders, but are also accompanied by side effects, both in short-term and chronic administration (4). GCs exert their influence on all tissues, including the immune system, where they acutely suppress the immune response (5). This immunosuppressive effect is particularly leveraged in the treatment of autoimmune disorders. On the other hand, chronic stress has been associated with a decrease in the function of this inhibition and is often associated with a pro-inflammatory profile, a risk factor for a number of medical and psychiatric disorders (6). However, large inter-individual differences are reported in the consequences of acute or chronic stress exposure as well as side effects experienced from GCs.

GCs exert their effects via mineralo- and glucocorticoid receptors (GRs), which are types of nuclear steroid hormone receptors. These receptors directly bind to gene regulatory elements at the DNA level and can either stimulate or repress gene transcription, leading to local epigenetic alterations (7). Individual variations in response to GC exposure can be mediated in part by altered effects at the level of gene regulation. Using massively parallel reporter assays in cell lines and stimulated expression quantitative trait locus (eQTL) analyzes in peripheral blood, we have previously reported that genetic variants can alter the effects of GCs at gene regulatory elements (8). These variants are linked to a spectrum of outcomes, including altered risk for psychiatric disorders, variation in amygdala reactivity to threat, startle response, and cortisol response to a psychological stressor (8–10) stress-related changes in brain physiology, such as the peak latency of the hemodynamic response function in limbic brain regions (11). In addition to genetic variation, the activity of glucocorticoid response elements (GREs) is also influenced by the local epigenetic landscape, including at the level of DNA methylation (DNAm) (12,13). Furthermore, it has been shown that GR-activation can lead to changes in DNAm at its direct binding sites, mainly through DNA demethylation (14–16). Beyond the effects of transcription factors on chromatin accessibility and DNAm at gene regulatory elements (17), which are likely main mediators of the extensively explored environmental influences on DNAm (18), DNAm patterns can also be influenced by genetic variants known as methylation quantitative trait loci (meQTLs). A recent study highlighted the association between DNAm and single nucleotide polymorphisms (SNPs) in a large meta-analysis, mapping the role of meQTLs in shaping the epigenome (19). Furthermore, tissue-specific meQTLs have been identified (20), and studies focusing on cell-type-specific meQTLs have provided insights into the cell-specific effects of genetic variants on DNAm patterns (21,22). Besides influencing baseline DNAm levels, genetic variation may also alter the above-described impact of GR activation on DNAm in various tissues (15,23). However, to date, no study has systematically explored how common genetic variation can moderate this impact, which will be the primary focus of this manuscript.

As indicated above, the regulation of genes by GCs, and consequently its impact on disease risk or treatment outcomes, is multifactorial, involving genetic, epigenetic, and transcriptional factors. Multi-omics approaches have emerged as powerful tools to disentangle such complex regulation (24), allowing for a comprehensive investigation of the molecular landscape that contributes to disease risk as these different levels are interrelated (25,26). We, for example, have shown that lasting GR-activation induced alterations in DNAm within gene regulatory regions can influence the subsequent effects of GR-activation on gene expression levels (19). This supports that changes in DNAm in response to stress-related stimuli can impact cellular responses and potentially contribute to the pathogenesis of various diseases.

In this study, we conducted the first multi-omics analysis investigating the genetic moderation of the effects of GR-activation via the agonist dexamethasone on changes in DNAm and gene expression. By integrating stimulus-dependent molecular QTLs into a comprehensive network analysis, we jointly analyzed DNAm and gene expression in response to GR-activation in the context of common genetic variations. This analysis revealed a multi-omics signature associated with GR-induced responses in peripheral blood as well as novel connections between functional, molecular GC-effect modulating variants and disease risk variants.

## Methods and Material

### Study samples

The study involved 202 participants from the Max Planck Institute of Psychiatry (MPIP), consisting of 68 women and 134 men. This included 88 individuals treated for major depressive disorder and 114 healthy controls (dataset GSE64930). Baseline blood samples were collected following specific protocols (see Supplementary Information), and participants were administered 1.5 mg dexamethasone orally at 6 pm with a second blood sample taken three hours post-ingestion. The study was approved by the ethics board of the Ludwig Maximilians University (approval #244/01) and was conducted in accordance with the current version of the Declaration of Helsinki.

### DNA methylation profiling

DNAm in our study was evaluated using Illumina EPIC v1 Methylation arrays, with a total of 404 blood samples analyzed, comprising 202 samples for each of the two time points. Data processing involved several steps detailed in Supplemental Information, leading to a final dataset comprising 740,357 CpGs across 398 samples.

### Differential DNA methylation analysis

To assess GR-induced alterations in DNA methylation, we utilized a linear mixed-effects model (R package lme4) on filtered, normalized, and batch-corrected methylation beta-values, both before and after dexamethasone treatment. This analysis accounted for covariates such as sex, age, BMI, depression status, estimated white blood cell counts, and the first two principal components of the genotype data, capturing the genetic variability of our study group. Differentially methylated positions (DMPs) were identified using a false discovery rate (FDR) threshold of <0.05.

### Genotype data and imputation

Human DNA from EDTA blood samples was genotyped using Illumina Human610-Quad (n = 79) and OmniExpress (n = 120) BeadChips, followed by quality control and imputation as previously described (27). The final dataset included 5,617,712 SNPs across 199 samples.

### Methylation quantitative trait loci analysis

The GR-meQTL analysis focused on evaluating SNP-CpG pairs within a ±1Mb region, utilizing standardized methylation changes (differences in DNAm post-dexamethasone treatment relative to baseline, standardized against the baseline). This analysis was conducted using the R package MatrixEQTL (28). We identified significant baseline and GR-meQTLs at an FDR < 0.05. For a more comprehensive description of the methodology, please refer to the Supplemental Information.

### Gene expression data

Baseline and GR-induced gene expression in blood from 199 individuals was assessed using Illumina HumanHT-12 BeadChips (GSE64930 and (27)). This data underwent quality control (see Supplemental Information), resulting in a dataset encompassing 11,944 transcripts from 398 samples.

### Expression quantitative trait methylation analysis

The GR expression quantitative trait methylation (eQTM) analysis encompassed 11,944 transcripts and 740,357 CpG sites, utilizing standardized changes in expression and methylation across a 2Mb window. Data analysis was carried out using MatrixEQTL, with covariate adjustment similar to the differential DNAm analysis and meQTL analyses. This included incorporating Surrogate Variables (SVs) 1-3, derived from the gene expression data to account for unobserved confounding factors. Significant *cis* eQTMs, which denote CpG-transcript pairs within a ±1Mb region, were identified using FDR < 0.05.

### Functional genomic annotation and characterisation

Differentially methylated CpGs and GR-meQTLs were annotated for genomic features using the minfi R package and UCSC genomic data (29). MeQTLs were additionally annotated for GR-binding sites, and chromatin state enrichment was assessed using ChromHMM annotation (30,31). For a detailed description of these analyses, see Supplemental Information.

### Multi-omics network inference and analysis

Multi-omics network inference and analysis were performed using KiMONo (32), which incorporated standardized changes in expression and methylation, along with genetic variants and established relationships. Detailed methodologies and the parameters used for evaluating network robustness are available in the Supplemental Information.

### GO enrichment analysis

For the GO enrichment analysis, we employed FUMA GENE2FUNC (33) with default parameters. The background list consisted of the 11,994 transcripts from our dataset.

### Gene Overlap with genes induced by GR activation in the mouse brain

To compare GR-network trios QTL genes, specifically SNP-CpG-transcript combinations where meQTLs function as both eQTLs and eQTMs, we utilized GR-response genes in mouse brain tissue data from DiffBrainNet (34). Orthologs were mapped by using the R package orthologsBioMART. The significance of overlap was determined using Fisher’s exact test in the R package GeneOverlap.

### GWAS enrichment analysis

For our GWAS enrichment analysis, we matched LD-independent GR-trio SNPs to GWAS variants based on chromosome and position coordinates (hg19). These SNPs were identified through a clumping process using PLINK (v1.90b5.3); details are provided in the Supplemental Information. Our analysis covered various psychiatric disorders and traits, cytokines, metabolic markers, and autoimmune diseases, using GWAS summary statistics with nominal p-value cutoffs (further details in the Supplemental Information). To create a background dataset for our analysis, we incorporated GR-meQTL (me)SNPs that were not included in the GR-trio set. We then conducted an enrichment analysis using 1,000 permutations to assess the significance of our findings. This evaluated the overlap with GWAS data, from which we calculated empirical p-values and odds ratios.

## Results

In this study, we conducted a multi-omics analysis in 199 individuals, including 85 patients with Major Depressive Disorder (MDD) and 114 healthy controls. We assessed mRNA expression and DNAm in peripheral blood cells, both at baseline and 3 hours post dexamethasone administration. Our comprehensive analysis included approximately 740k CpG sites, 5.62 million SNPs, and 12k transcripts (Figure 1a).

**Figure 1:**
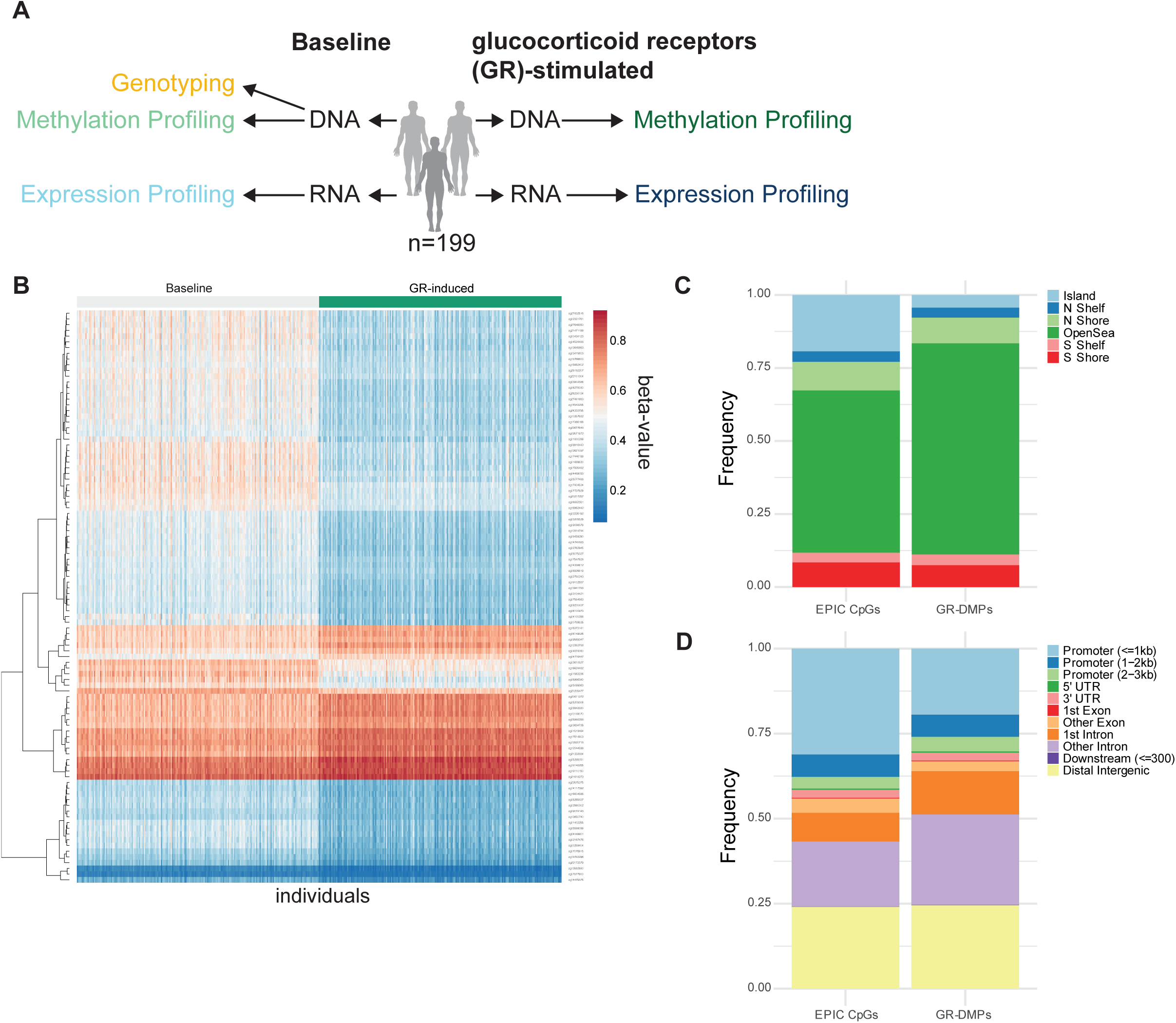
Overview of the GR-meQTL Study. (**A**) The stepwise experimental design used to investigate the genetic effects on the glucocorticoid receptor (GR) response in human whole blood involves several key steps: (1) Treatment of 199 individuals with 1.5 mg dexamethasone per os. (2) Transcriptome and methylome measurements were obtained from the entire cohort at two time points: baseline and 3 hours post-treatment. This included DNAm patterns assessed using the Illumina MethylationEPIC BeadChip, aligning with the time-point of mRNA expression measures (same blood draw with Paxgene RNA and EDTA tubes). (3) Genotype profiling was performed, utilizing a subset of our previously published GR-eQTL analysis, to map GR-meQTLs. (4) eQTM analysis was conducted to explore the relationships between gene expression and methylation. (5) Multi-omics network inference and analysis were carried out, integrating the data to understand the genetic impact on GR responses. (**B**) Mean methylation profiles of differentially methylated positions (DMPs) (n=3,280, FDR 5%) in 199 individuals, illustrating changes between baseline and GR-induced conditions. The top 100 DMPs are depicted. (**C-D**) Annotation of GR DMPs: (**C**) CpG islands; N(S) Shore, 2Kb-long regions flanking both sides of a CpG island; N(S) Shelf, 2Kb upstream/downstream of the furthest limits of the CpG shores, excluding CpG islands and CpG shores; OpenSea, encompassing the remaining genomic regions and (**D**) gene locations are indicated for a comprehensive understanding of the genomic context of the identified DMPs.

### Uncovering in vivo GR-induced differential DNA methylation

We conducted a differential DNAm analysis to examine the effects of dexamethasone on DNAm in peripheral blood cells, accounting for case-control status, sex, age, BMI, estimated white blood cell subpopulations, and the first two principal components derived from genotype data. Applying a 5% FDR correction, we identified 3,280 CpG sites showing significant methylation changes after GR-activation (GR-DMPs, Table S1, Figure 1b), with an average absolute DNAm difference of 9.2% (ranging from −17.5% to 12.9%). Among the 3,280 GR-DMPs, 76.4% (2,506 CpGs) were hypomethylated, and 23.6% (774 CpGs) were hypermethylated. The majority of GR-DMPs were located outside of CpG islands (open seas) (72.3%), distal intergenic (24.5%), and intronic regions (39.3%), differing from the EPIC array’s general distribution, where 55.5% are in open seas, 27.6% in introns, and 41.2% in promoter regions (Fig. 1c and Fig. 1d).

### Overlapping as well as distinct mechanisms of genetic control: contrasting GR-meQTLs with baseline meQTLs

To explore the genetic regulation of GR-induced DNAm changes, we carried out a *cis*-meQTL analysis to assess associations between SNPs and DNAm changes at CpG sites within a 1Mb window. We identified 104,828 significant GR-meQTLs, involving 3,772 unique CpGs and 88,585 unique SNPs, with a small overlap of 12 GR-meCpGs with GR-DMPs (Table S2, Figure 2a). The average distance between meSNPs and meCpGs was 105 Kbp. Additionally, we performed meQTL analysis using baseline methylation levels (see Table S3 for details). Among the GR-meCpGs, 69% overlapped with baseline meCpGs (n=2,618 CpGs). Examples of GR-meQTLs with and without a baseline meQTL are illustrated in Figure 2b-c. We further characterized the identified GR-meCpGs and GR-meSNPs in terms of their genomic location and regulatory features. Among the 3,772 GR-meCpGs, only 16.7% were located in CpG islands, while 23.2% were in CpG island-adjacent regions (shores and shelves). The majority (60.1%) were located in open seas (see Fig. 2d). In terms of regulatory features, 37.7% of GR-meCpGs and 10.9% of GR-meSNPs were located in promoters, while a substantial proportion of GR-meSNPs resided in introns (54.6%). Additionally, 25.6% of GR-meCpGs and 32.7% of GR-meSNPs were located in distal intergenic regions (see Fig. 2e-f). When compared to baseline analyses, GR-meSNPs were more often located in introns (53.6% vs. 46.3%) and less often in distal intergenic regions (32.7% vs. 43%) (refer to Fig. 2d-f). Moreover, GR-meCpGs were more frequently located within GREs compared to baseline meCpGs (25.1% vs. 20.3%, refer to Fig. 2g-h). Validation of baseline meQTLs using Genetics of DNA Methylation Consortium (GoDMC) data showed a 77.6% replication rate (see Supplementary Information).

**Figure 2:**
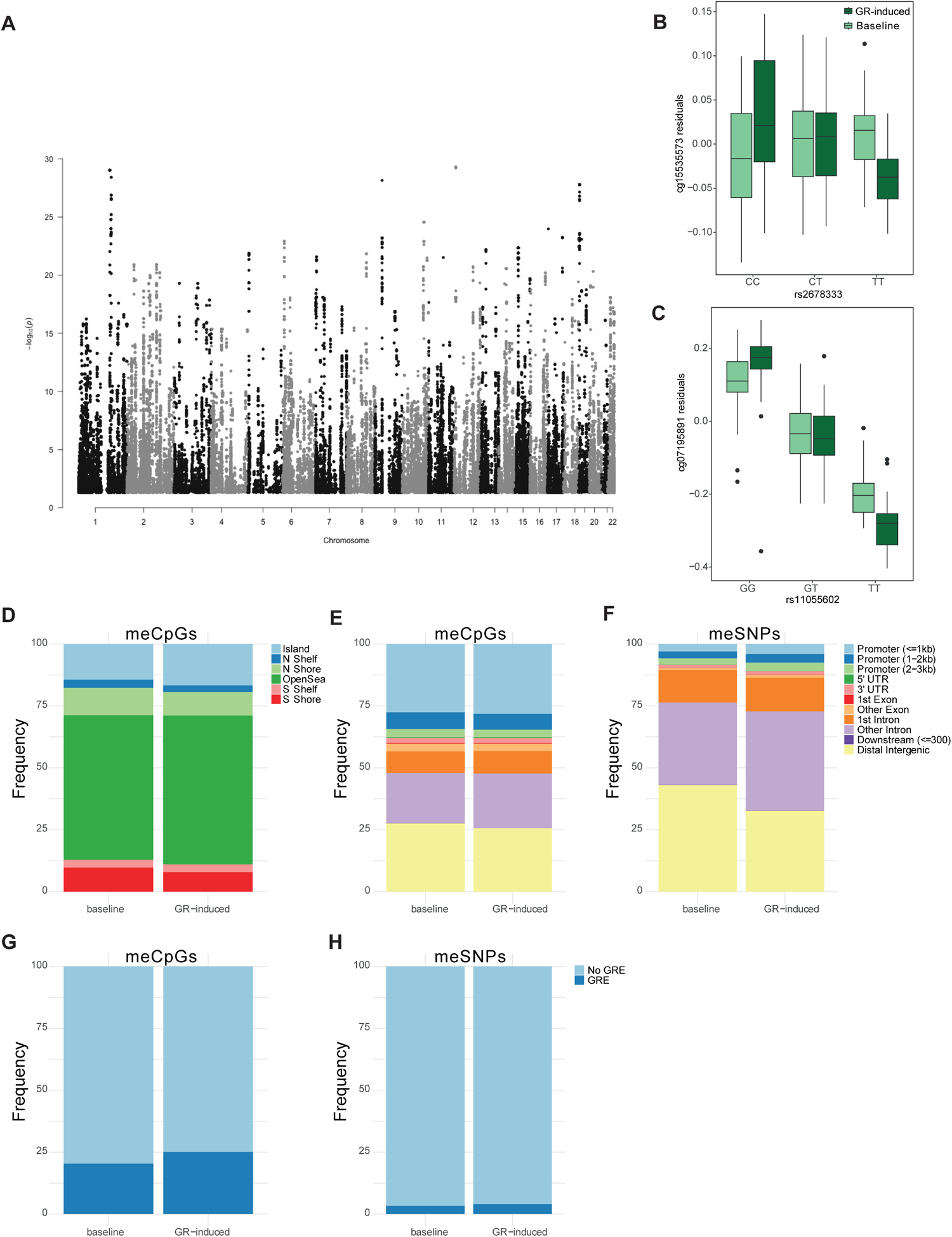
GR-meQTL Analysis. (**A**) Manhattan plot displaying the results of the GR-meQTL analysis, illustrating the distribution of 104,828 local GR-meQTLs (88,585 unique GR-meSNPs) across the genome. (**B-C**) Boxplots showing the residualized beta methylation values of significant GR-meQTLs as examples. Methylation levels are stratified based on the genotypes of the meSNPs. (**B**) A GR-meQTL (rs2678333-cg15535573) located on chromosome 12 with a post-dexamethasone-specific effect (FDR = 8.24×10^-15^). (**C**) A GR-response meQTL (rs11055602-cg07195891) located on chromosome 12, exhibiting effects both before and after dexamethasone treatment, with a notable decrease in individuals with major genotype and an increase in individuals with the minor allele genotype in post-dexamethasone in cambrian to baseline (FDR = 5.1×10^-30^). (**D-E**) Genomic characteristics of GR-meCpGs and baseline meCpGs in relation to (**D**) CpG islands and (**E**) nearby genes. Various genomic regions were analyzed: CpG islands; N(S) Shore, 2Kb-long regions flanking both sides of a CpG island; N(S) Shelf, 2Kb upstream/downstream of the furthest limits of the CpG shores, excluding CpG islands and CpG shores; OpenSea, encompassing the remaining genomic regions. (**F**) Characteristics of GR-meSNPs are detailed in relation to the genomic location of nearby genes. (**G-H**) GR- and baseline-meCpGs and meSNPs annotated for glucocorticoid response element (GRE) proximity.

### Consistent regulatory patterns of GR-meQTLs across blood cell types

To understand the functional implications of GR-meQTLs, we examined their distribution across functional genomic regions using ChromHMM epigenetic states in blood and T-& B-cell lines. Our analysis indicated that GR-meCpGs were more frequently found in promoters and enhancers compared to GR-meSNPs (see Fig. 3a-b). Additionally, we found that 44.6% of GR-meCpG sites were associated with SNPs localized to the same chromatin state. Interestingly, neither GR-meCpGs nor meSNPs showed significant differences in the distribution of epigenetic states across single blood cell types (pairwise Wilcoxon p-value >0.05, Figure 3a-b).

**Figure 3:**
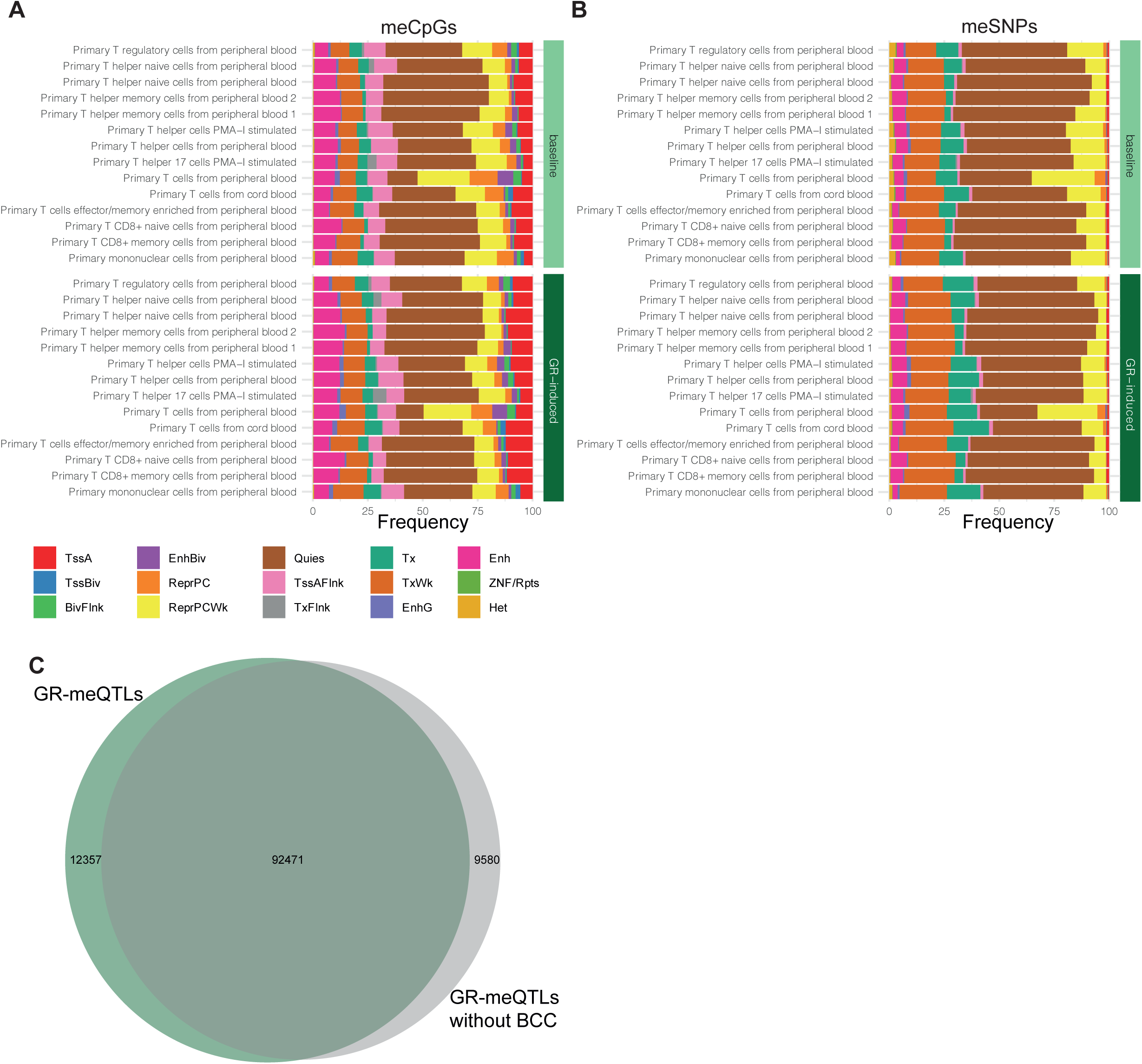
GR-meQTLs and Blood Cell Types. (**A-B**) This shows histone mark enrichment for GR-meCpGs (**A**) and GR-meSNPs (**B**) across 15 chromatin states in 18 primary blood cell types. The top panel illustrates the distribution in baseline meQTLs, with GR-meCpGs being more frequent in promoters (17.6% vs. 2.3% in GR-meSNPs) and enhancers (14.8% vs. 6.9% in GR-meSNPs). The bottom panel focuses on GR-meQTLs, noting that 44.6% of meCpG sites are associated with SNPs in the same chromatin state but different positions. Analysed chromatin states include TssA (Active TSS), TssAFlnk (Flanking Active TSS), TxFlnk (Transcr. at gene 5’ and 3’), Tx (Strong transcription), TxWk (Weak transcription), EnhG (Genic enhancers), Enh (Enhancers), ZNF/Rpts (ZNF genes & repeats), Het (Heterochromatin), TssBiv (Bivalent/Poised TSS), BivFlnk (Flanking Bivalent TSS/Enh), EnhBiv (Bivalent Enhancer), ReprPC (Repressed PolyComb), ReprPCWk (Weak Repressed PolyComb), and Quies (Quiescent/Low). (**C**) A Venn diagram highlights the overlap between GR-meQTLs adjusted for white blood cell counts and those without adjustment. 102,051 meQTLs (86,834 meSNPs and 3,839 meCpGs) were identified without BCCs adjustment, of which 90,6% (n=92,471 meQTLs) were found to be common with the original GR-meQTL analysis that accounted for white BCCs.

Since cell type-specific meQTLs have been reported, we further explored the cell type-specificity of GR-meCpGs by re-analyzing GR-meQTLs without adjusting for blood cell compositions. This revealed 102,051 meQTLs, of which 90.6% overlapped with those identified in the original analysis that included white blood cell compositions (Fig. 3c). These findings suggest that GR-meQTLs do not show strong cell type-specific effects in peripheral blood.

### Absence of shared genetic control: No overlap between GR-induced DNAm and GR-induced gene expression changes

To investigate the genetic control of gene expression and DNAm, we compared the patterns of GR-meQTLs and GR-eQTLs. By leveraging our previously identified *cis* GR-eQTL results (27), with 717 GR-eQTL transcripts and 10,078 GR-eQTL(e)SNPs across 297 individuals, we discovered a total of 9,688 non-overlapping eSNPs. Strikingly, a significant proportion (96%) of these eSNPs was exclusively detected by GR-eQTL analysis and not in the GR-meQTL analysis (see Fig. 4a). Among the GR-meSNPs, only 0.44% (n=390) were also identified as GR-eSNPs impacting 591 meQTLs. In contrast, our baseline analysis revealed a substantial overlap of 89% (n=150,057) between our previously reported baseline eSNPs (n=167,885) and meSNPs (n=2,274,829).

**Figure 4:**
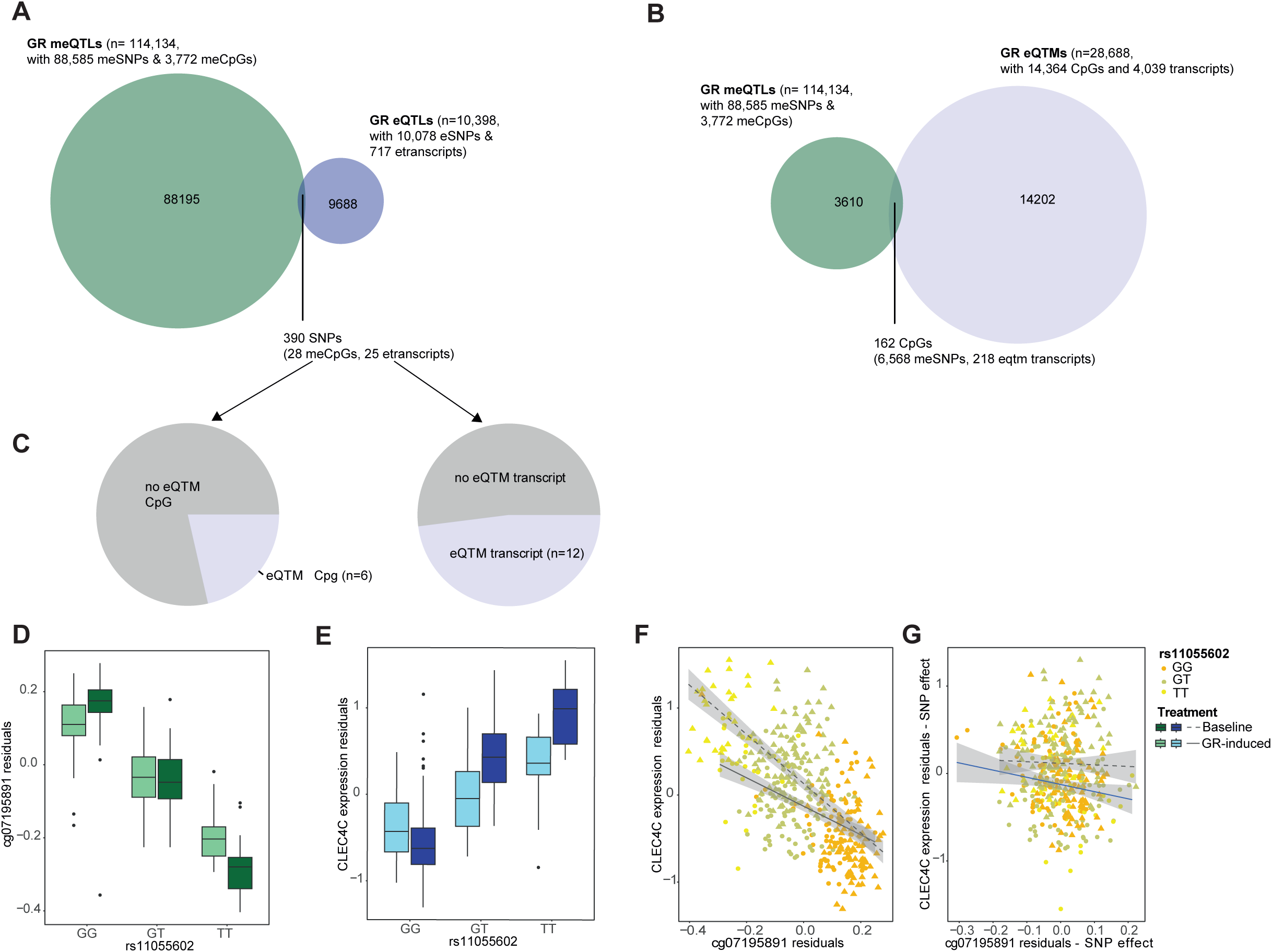
Integrative Multi-omics Analysis. (**A**) Venn diagram representing the intersection of GR-meSNPs (methylation SNPs) and GR-eSNPs (expression SNPs). Our analysis uncovered 591 GR-meQTLs, including 390 SNPs and 28 CpGs, that share SNPs with GR-eQTLs. (**B**) Venn diagram representing the intersection of GR-meCPG (methylation CpG sites) and GR-eQTM (expression quantitative trait methylation) CpG sites. (**C**) Pie chart showing the distribution of eQTM transcripts and eQTM CpG sites that overlap with the CpGs and transcripts from the shared GR-meSNPs and GR-eSNPs overlap. (**D-E**) Boxplots displaying the residualised beta methylation values and expression levels for the GR-trio hit *CLEC4C*, stratified by SNP rs11055602, with this locus acting as an eQTL, eQTM, and meQTL. (**F**) Scatterplot illustrating the relationship between residualised methylation and expression, highlighting the SNP effect. (**G**) Scatterplot showing the relationship after adjusting for the SNP effect, indicating that the correlation at baseline disappears post-adjustment.

### Large number of associations between GR-induced DNAm and GR-induced gene expression

To explore the relationship between GR-induced changes in DNAm and changes in gene expression, we conducted an eQTM analysis using the gene expression data from our sample. Employing the same analytic approach as for GR-meQTLs, we identified a substantial number of 28,688 *cis* GR-eQTM associations (consisting of 14,364 GR-eQTM CpGs and 4,039 GR-eQTM transcripts) in our dataset at an FDR of 5% (Table S4). However, only a small proportion of 162 GR-eQTM CpGs overlapped with our GR-meCpG sites (see Fig 4b), suggesting that while GR-response mechanisms exhibit correlation at both DNAm and gene expression levels, only a small fraction (4%) is under genetic influence. It is noteworthy that we identified merely 1,307 eQTMs at baseline (consisting of 1,126 eQTM CpGs and 512 eQTM transcripts, see Table S5).

### Integration of GR-me/eQTLs and GR-eQTMs via multi-omics network analysis: Capturing immune-related pathways

We conducted an overlap analysis to identify common GR-induced loci by comparing the physical positions of GR-meQTLs with significant pairs from GR-eQTLs and GR-eQTMs. This analysis revealed a limited overlap (Fig. 4a), with more details provided in the Supplementary Information. A notable example is the *CLEC4C* gene, which demonstrated multiple omics associations and significant eQTMs correlations both at baseline and after GR-stimulation (Fig. 4b-c). However, when adjusting for me/eSNP effect, the baseline correlation disappeared (Fig. 4c). This finding led to a more comprehensive investigation, as it suggested the potential for false positive me- or eQTLs. Such biases can only be identified through a multi-omic analysis. Consequently, we generated an integrated multi-omic network using KiMONo (32), combining methylation, transcriptomic, genetic data and biological variables like sex, age, BMI, and case-control status. The resulting GR-network, with 7,193 nodes and 30,332 edges, reflects the complex interplay between transcripts, SNPs, CpGs, and biological factors. Stringent criteria were applied to ensure network robustness (see Supplementary Information and Table S6).

We further examined GR-network trios (Fig. 5a), which are GR-meQTLs also acting as GR-eQTLs and GR-eQTMs, thus forming SNP-CpG-transcript combinations or trios. Our analysis revealed 552 eQTMs and 297 eQTLs, mapping back to 7,979 GR-meQTLs (Fig. 5b and Table S7). Among these, we discovered 7,591 GR-trio SNPs, 334 GR-trio CpGs, and 613 GR-trio genes. These trio genes showed significant enrichment in immune-related pathways (see Fig. 5c), and a significant overlap with genes regulated by dexamethasone in the mouse brain (Odds Ratio: 1.8, p-value: 1.4×10^-6^, Fig. 5d and Table S8) (34). Interestingly, 59% of these genes were differentially regulated in the prefrontal cortex (Fig 5e). Additional details are available in Supplementary Information. These findings highlight the pivotal role of these trio genes in stress impacts on the brain and their critical influence in mediating glucocorticoid effects on brain function and behavior.

**Figure 5:**
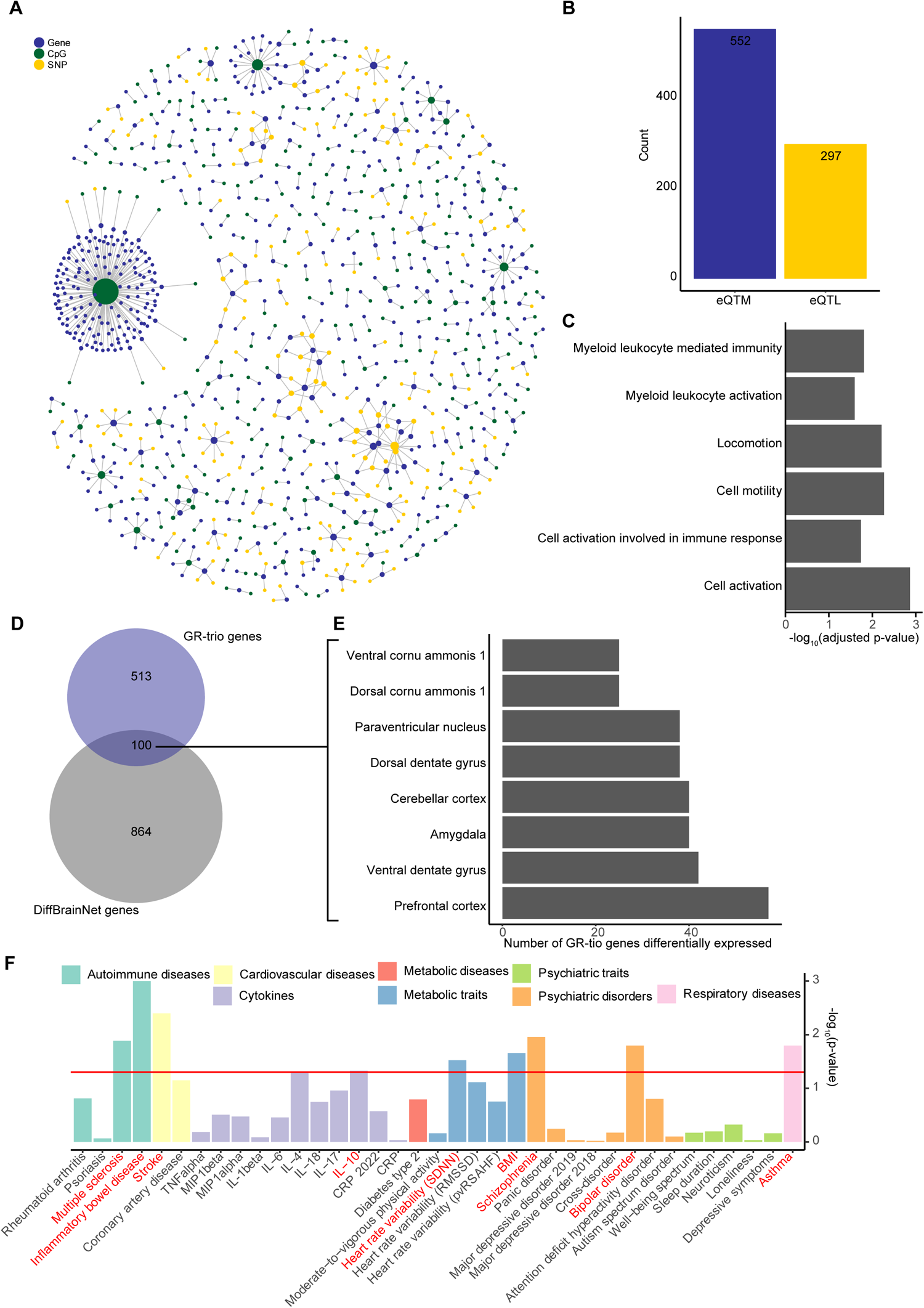
**GR-trios: (A**) The GR-trio network, composed of meQTLs acting as both eQTLs and eQTMs, forming SNP-CpG-transcript trios. The network is color-coded, with SNPs in yellow, genes in blue, and CpGs in green. (**B**) A barplot illustrating the distribution of eQTM and eQTL relations within the GR-trio network. (**C**) The GR-trio genes displayed enrichment in immune-related GO pathways such as myeloid leukocyte-mediated immunity, cell activation involved in immune response, and myeloid leukocyte activation, as well as pathways related to cell activation, cell motility, and locomotion. (**D**) Venn diagram displaying the overlap between GR-trio genes and genes differentially regulated in the mouse brain following dexamethasone administration. (**E**) A bar plot depicting the distribution of the overlapping genes across various brain regions. (**F**) Enrichment of nominal GWAS associations for the GR-trio SNPs across various GWAS datasets, including psychiatric disorders, psychiatric traits, cytokines, metabolic markers, autoimmune diseases, and asthma. The Y-axis represents the enrichment p-value compared to non-trio GR-meSNPs, with significant enrichment indicated by red labels. Bars are color-coded according to their disease or trait class.

A key hub within this network was identified as a single CpG site (cg09614808) in the *Amyloid precursor protein* (*APP*) gene, connected with 164 genes across all 22 autosomes. Additional notable hubs included cg09614808 in the *Tyrosine 3-Monooxygenase* (*YWHAZ*) gene and cg17764313 in the *Minichromosome Maintenance Deficient 2* (*MCM2*) gene, connected to 14 and 25 genes, respectively.

### The GR-induced SNP-methylation-mRNA trios are relevant to psychiatric-related diseases

Given the established impact of stress on various health conditions, particularly in inflammation, autoimmune disorder, and psychiatric traits, we investigate whether GR-trio variants (7,591 SNPs, including 321 LD independent SNPs, Table S9), which modulate DNAm and gene expression in response to GR-activation also play a role in these conditions. Our GWAS enrichment analysis, using 80,994 non-trio meSNPs (including 3,267 LD independent SNPs) as a control group and considering summary statistics from diverse sources covering psychiatric disorders, psychiatric traits, cytokines, metabolic markers, autoimmune diseases, and asthma (detailed in the Methods section and Supplementary Information), showed significant enrichment of GR-trio SNPs in several conditions. This included bipolar disorder (odds ratio [OR] = 1.36, permutation p-value [p] = 0.016), schizophrenia (OR = 1.38, p = 0.011), multiple sclerosis (OR = 1.45, p = 0.013), asthma (OR = 1.32, p = 0.016), stroke (OR = 1.51, p = 0.004), BMI (OR = 1.67, p = 0.022), inflammatory bowel disease (OR = 1.77, p < 0.001), IL-10 (OR = 1.41, p = 0.047) and heart rate variability (OR = 1.78, p = 0.03), see Fig. 5f.

## Discussion

In this study, we used advanced network analysis and multi-level GR-response data to explore 1) the influence of genetic variants on DNAm patterns in response to GC exposure, 2) the consequent effects of these changes on gene expression when induced by GCs, and 3) the associated genetic risk for various medical diseases and traits. While recent investigations have begun exploring multi-omics data in various fields (for example (35–38)), however none have yet integrated data that reflect dynamic changes following a disease-relevant stimulus. Here, we leveraged such data with repeated measures before and after GC stimulation in 199 individuals, integrating across genetic, DNAm, and gene transcription data. Our network model incorporated 88,585 GR-meSNPs and 3,772 GR-meCpGs, along with approximately 12,000 transcripts. This set was embedded in the context of all array CpG sites (∼800k CpGs), and our ∼5.6 million imputed SNPs. Our analysis highlights not only the dynamic changes in DNAm following GR-activation, but also the different levels of regulations with distinct effects of genetic variants on changes in GC-induced gene expression and DNAm. Focusing on GR-trio variants from our multi-omics model, we showed that genetic variants that alter the molecular impact of GCs in immune cells are associated with risk for psychiatric disorders, metabolic traits, autoimmune disorders, and cardiovascular disease as well as related traits. This supports the observations that chronic stress, partly mediated by GC exposure, is a shared risk factor across psychiatric and medical diseases. Our findings suggest that the genetic variants moderating the effects of GCs, as identified, may underlie some of the genetic correlations observed across these traits (for example (39–41)).

In our study, we conducted a differential analysis of GR-induced DNAm to evaluate the primary impacts of GR on DNAm. We identified 3,280 CpG sites displaying significant differential DNAm, with a substantial majority showing hypomethylation (76.4%), mainly situated in open sea regions. This finding aligns with existing reports, such as (16), suggesting that GR-binding can lead to active DNA demethylation through active DNA-repair mechanisms. Furthermore, it concurs with a recent insight from (17), which indicates similar mechanisms for local epigenetic changes following transcription factor binding in general.

While we discovered a substantial number of GR-meQTLs (N=104,828), these exhibited distinct patterns and mechanisms compared to previously identified GR-eQTLs (27), with less than 1% overlap at the SNP level. Discrepant levels of overlap, ranging from substantial (42) to less than 5% (43), have previously been reported, likely highlighting the contextual nature of the interaction between meQTLs and eQTLs. The cell type specificity of baseline DNAm, even within blood cell types, is well-established (44). In contrast to this pattern, our study found that only a smaller subset of GR-meQTLs displayed such specificity. This suggests that epigenetic modifications related to stress may have shared genetic regulation across a diverse range of immune cell types and possibly even beyond the immune system. Moreover, the transcripts within the GR-trio genes from our multi-omics analysis were responsive to GCs in various mouse brain regions (34). This observation aligns well with previous findings indicating that GR-responsive sequences, as identified in eQTL and massively parallel reporter assays, are enriched in cross-tissue enhancers (8,10). Consequently, the shared genetic associations observed in the GR-trio SNPs, spanning a range of psychiatric and medical disorders and physiological traits, may extend their role in moderating stress effects in the immune system to possibly influence other tissues, including the brain.

Additionally, we performed an eQTM analysis specifically related to GR-activation. This analysis revealed correlations between GR-induced changes in both gene expression and methylation at a larger number of CpG sites (N=14,364). Remarkably, only a small fraction (4%) of these CpG sites was regulated by GR-meQTLs, indicating that a proportion of molecular GR effects are not strongly moderated by genetic variation. This observation aligns with the physiological importance of these effects in a coordinated stress response. However, some GR-eQTM outcomes were influenced by genetic variants. For instance, the *CLEC4C* locus, which encodes C-Type Lectin Domain Family 4 Member C, shows its dual role in both GR-e/meQTL and GR-eQTM associations. Notably, upon adjusting for eSNP and meSNP effects, the correlation strength altered, persisting only when stimulated (Figure 4). This emphasizes the critical role of genetic factors in influencing GR-responses in DNAm and gene expression, highlighting the necessity of integrating multiple omics layers to avoid false positives and negatives.

While our study contributes significant insights, there are limitations that should be considered. We focused on bulk RNA and DNA measures from peripheral blood cells, and future studies should incorporate high-resolution methodologies at single-cell resolution for a more comprehensive understanding of cellular heterogeneity, recently highlighted in the human brain cell atlas (45). Additionally, investigating different exposure lengths and timings of GR-stimulation may provide further insights into the temporal dynamics of the molecular response. While trans effects are considered in our multi-omic network analysis, it should be noted that our GR-meQTL analysis primarily focuses on *cis* meQTLs. Therefore, network nodes with many trans-effects, such as the CpG site within APP, should be interpreted cautiously and will need replication.

By generating multi-omic networks of the regulation of the molecular response to GC in immune cells, we identified shared genetic influences of “stress” moderating variants on a range of diseases - psychiatric to medical - that have previously been reported to be affected by chronic stress and GC exposure. By pinpointing key multi-level regulators, we enhance the patho-mechanistic understanding of how stress can serve as a transdiagnostic risk factor. This, in turn, paves the way for the identification of potential diagnostic and therapeutic targets.

## Data and Code availability

All computational code has been made available on GitHub: https://github.com/jArloth/GR-meQTLs/, while the DNA methylation data are accessible in the GEO repository under GEO: GSE249113.

## Acknowledgments

We extend our gratitude to Linda Dieckmann, Darina Czamara, and the medical genomics group for their invaluable insights and discussions throughout the project’s execution. Our heartfelt appreciation goes to all the participants whose involvement was essential to the success of this study. The authors thank OpenAI for providing assistance through ChatGPT during the writing process of this manuscript. This work was supported by European Union’s Horizon 2020 Research and Innovation Programme under Grant Agreement No 860895, European Training Network grant, Translational SYStemics: Personalised Medicine at the Interface of Translational Research and Systems Medicine (TranSYS), and European Research Council (ERC) starting grant number 281338, “Gene x environment interactions in affective disorders – elucidating molecular mechanisms” (GxE molmech) to Elisabeth Binder. Dr. Knauer-Arloth’s contributions were supported by the Brain & Behavior Research Foundation (NARSAD Young Investigator Grant, No 28063).

## Contributions

J.K.-A. conducted the computational analysis, generated data visualizations, and drafted the initial manuscript. E.B.B. contributed intellectually, conducted substantial manuscript revisions, and provided critical feedback as well as funding. J.K.-A. and E.B.B. collaboratively conceptualized, designed, and supervised the study. A.H. was responsible for the quality control of the methylation data. All authors had the opportunity to review, offer feedback on, and approve the final manuscript.

