## Supplemental Information for "Multi-omics analysis of the molecular response to glucocorticoids - insights into shared genetic risk from psychiatric to medical disorders"

### Supplementary Methods and Material

#### Detailed Study Samples Description

The study involved 202 participants from the Max Planck Institute of Psychiatry (MPIP), consisting of 68 women and 134 men. Among these, 88 individuals were receiving treatment for major depressive disorder at MPIP's hospital, while the remaining 114 were healthy controls without a history of psychiatric disorder. The genotype and gene expression data of this cohort represents a subset of the publicly available dataset GSE64930. Baseline whole blood samples were collected at 6 pm after a two-hour fasting period and abstention from coffee and physical activity. Subsequently, participants were administered 1.5 mg dexamethasone per os, and a second blood sample was obtained at 9 pm, three hours after dexamethasone ingestion. Detailed recruitment strategies and cohort characterization are described in previous works (1,2).

#### Comprehensive DNA Methylation Profiling

Four hundred four whole blood samples (baseline and GR-induced) were run on Illumina EPIC v1 Methylation arrays. The arrays were randomized based on sex and case-control status, ensuring that both time points for each participant were kept together. All DNA methylation samples were processed using the pipeline described in (3). Scan intensity signals stored in .idat files were loaded into R and transformed into beta-values within the R package minfi (3,4). Samples with a mean detection p-value > 0.01 and those showing sex mismatches between estimated sex from methylation data and confirmed phenotypic sex were excluded (n=1). Beta-values were normalized using stratified quantile normalization followed by BMIQ (5). CpG probes on X or Y chromosomes, probes containing SNPs, cross-hybridizing and polymorphic probes (6), and probes with a detection p-value of > 0.01 in one or more samples were removed. Afterward, the beta values were transformed into M-values, and batch effects of plate, array, and slide were removed using Combat (7). For this, a principal component analysis (PCA) on the M-values was carried out. The association between batches and the principal components was checked iteratively until no significant batch was left. Corrected M-values were re-transformed into beta-values. The EpiDISH method was used to estimate blood cell type composition (8). Finally, the MixupMapper (9) was applied to both genotype and methylation data to check for possible sample mix-ups. Two mix-up individuals were found, and four samples were excluded from the further analysis. The final dataset contains 740,357 CpGs and 398 samples.

#### In-depth Methylation Quantitative Trait Loci (meQTL) Analysis

The GR methylation quantitative trait loci (meQTL) analysis focused on SNP-CpG pairs within a ±1Mb region. For each CpG-SNP pair, the SNP was coded as 0, 1, or 2 based on the frequency of the minor allele in this sample. To determine GR-meQTLs, a linear model assessed the standardized methylation change (post - pre /pre dexamethasone) using the R package MatrixEQTL (10). This model was adjusted for the same covariates as employed for differential DNA methylation analysis. To identify baseline cis-meQTLs, the model utilized pre-dexamethasone methylation levels. The significance threshold was set at FDR < 0.05 to identify significant meQTLs.

#### Replication of meQTLs

Due to the unavailability of a cohort for GR-meQTL association replication, we utilized publicly available data from the Genetics of DNA Methylation Consortium (GoDMC) (11) to validate baseline meQTLs. Among the subset of CpGs present on the 450k array used by GoDMC, 48,092 out of 61,968 baseline meCpGs (77.6%) were also reported by the GoDMC study as meCpGs with a p-value < 1x10^-8^.

#### Gene Expression Data Collection and Processing

Transcriptome-wide measurements of baseline and GR-induced gene expression in blood were obtained for all 199 individuals using Illumina HumanHT-12 version 3 and 4 BeadChips and were previously published (GSE64930 and (1)). In brief, we utilised 11,994 autosomal filtered expression array probes. Each probe underwent variance stabilization and normalization (12) and known batch effects were corrected using ComBat. Surrogate Variable Analysis (12) was employed to address potential confounding from cell proportions or unaccounted factors. The resultant dataset encompassed 11,944 transcripts and 398 samples.

#### Functional Genomic Annotation and Characterization Procedures

Differential DNA methylated positions were aligned with their genomic and gene locations based on Illumina's annotation using the minfi R package. For assessing CpG-gene relations and genomic regions, the distance from each CpG to its annotated genes was computed, and the nearest genes were determined. If a single CpG was mapped to multiple regions, it was assigned to each respective region.

We annotated the genomic features of CpG sites obtained from the differential DNA methylation analysis and meQTL analyses, encompassing CpGs located in CpG islands. This annotation was performed using the minfi R package according to Illumina's annotation. In addition, sequence-based attributes were annotated using UCSC's genomic annotation for the hg19 genome build through the utilization of the TxDb.Hsapiens.UCSC.hg19.knownGene and ChIPseeker Bioconductor R packages (13). Furthermore, we determined genomic attributes for meSNPs as well. To examine the co-localization of baseline and GR-meSNPs with GR-binding sites, we used the Encode NR3C1 ChIP-seq data from GM12878 LCLs treated with dexamethasone (accession: GSE45638). To determine whether GR-meQTL CpGs and SNPs are enriched in specific chromatin states compared to baseline meQTLs, we utilized the 15-state annotation from ChromHMM within primary blood cells (n=18 cell lines) (14,15). For each meQTL, the best SNP or CpG was selected based on its meQTL association p-value.

#### Multi-Omics Network Inference and Analysis Protocol

Multi-omics network inference and analysis were performed using KiMONo (16) based on the standardized changes in expression and methylation, as well as on genetic variants. In this methodology, network nodes symbolize diverse features, including genes, CpGs, SNPs, and biological variables. The connections between these nodes signify established relationships within the input data. For network inference, three distinct priors were incorporated to provide existing knowledge about the connections between transcriptomic, epigenomic, genetic, and biological data. These priors serve as a foundational guide, streamlining the complexity and enhancing the algorithm's efficiency. The first prior was constructed using the BioMart tool (17) to link genes with their respective proteins. For the second prior, we leveraged information from the BioGRID database (18), which contains data on protein-protein interactions. Additionally, a prior was generated to link the omic information to our biological data. To assess network node robustness, three parameters were utilized: 1) goodness of fit (R2<0.1), 2) absolute effects (beta < 0.04), and 3) stability selection scores lower than 100% (k=100 runs).

#### GWAS Enrichment Analysis Procedures

For GWAS enrichment analysis, we matched LD-independent GR-trio SNPs to GWAS variants based on chromosome and position (hg19). To identify LD-independent SNPs, we employed a clumping procedure using PLINK (v1.90b5.3) with the specific parameters: --clump-r2 0.2 --clump-p1 0.05 --clump-p2 1 --clump-kb 1000). We used GWAS summary statistics with a nominal p-value cutoff for various psychiatric disorders, including attention-deficit-hyperactivity disorder (19), autism spectrum disorder (20), bipolar disorder (21), major depressive disorder (22,23), schizophrenia (24), post-traumatic stress disorder (25), and the cross-disorder associations (26); psychiatric traits: well-being spectrum (27), sleep duration (28), neuroticism (29), loneliness (30) and depressive symptoms (31); cytokines including C-reactive protein (CRP) (32) (33), Interleukin (IL)-10, IL-17, IL-18, IL-1b, IL-4, IL-6, Macrophage Inflammatory Protein-1 (MIP1) alpha, MIP1 beta and Tumor necrosis factor (TNF)-alpha (33); metabolic markers including heart rate variability (HRV) peak-valley respiratory sinus arrhythmia (pvRSAHF), HRV root mean square of successive differences (RMSSD), HRV standard deviation of the normal-to-normal interval (SDNN) (34), moderate-to-vigorous physical activity (MVPA) (35) and BMI (36); autoimmune diseases, including rheumatoid arthritis (37), inflammatory bowel disease (38), psoriasis vulgaris (39) and multiple sclerosis (40) as well as other diseases in which immune dysregulation play an important roles like cardiovascular diseases including coronary artery disease (41) and stroke (42), respiratory diseases like asthma (43) and metabolic disease such as diabetes type 2 (44).

To establish a background dataset (n=80,994 SNPs), we considered GR-meSNPs not included in the GR-trio SNP set. To minimise bias, we created MAF bins for each SNP set using 0.05 increments from 0 to 1. We conducted an enrichment analysis using 1,000 permutations, assessing the overlap of randomly selected background SNPs of the same size as the GR-trio SNP set and their MAF bin distributions with the GWAS summary statistics. The empirical p-value was calculated based on how frequently one of the 1,000 overlaps exceeded the actual overlap of the GR-trio SNP set with the GWAS dataset, normalized by the number of permutations. Additionally, we calculated an odds ratio (OR) by dividing the actual overlap by the mean of the 1,000 overlaps from resampling.

### Supplementary Results

#### Overlap of GR-me/eQTLs and GR-eQTMs

To uncover the common set of GR-induced loci, we performed an overlap analysis between the physical positions of GR-meQTLs (meSNPs and meCpGs) with significant pairs identified from the GR-eQTLs (eSNPs and etranscripts) and GR-eQTMs (eQTM transcripts and eQTM CpGs). We found that out of the 591 GR-meQTLs (consisting of 390 SNPs and 28 CpGs) that share SNPs with GR-eQTLs, six GR-meCpGs were correlated with GR-induced expression of eQTM transcripts (with 12 eQTMs involving 11 transcripts). Additionally, 28 GR-etranscripts shared 12 transcripts with GR-eQTMs (see Fig. 4a). An example of a locus that acts as an eQTL (rs11055602-CLEC4C), eQTM (cg07195891-CLEC4C), and meQTL (rs11055602-cg07195891) is the *C-Type Lectin Domain Family 4 Member C gene* (*CLECE4*) (Fig. 4b-c). In our previous analyses, we had identified *CLECE4* as one of our GR-eQTLs mapping to GWAS hits for major depressive disorder and schizophrenia. This gene plays diverse roles in cell adhesion, cell-cell signalling, glycoprotein turnover, inflammation, and immune response and exemplifies the importance of integrating all three omics data simultaneously. The eQTMs exhibit significant correlation at baseline and after GR-stimulation, but when adjusting for the SNP effects (rs11055602), the correlation at baseline disappears (Fig. 4c).

#### Integration of GR-me/eQTLs and GR-eQTMs with KiMONo

Given these findings, we proceeded to generate a fully integrated map by combining methylation, transcriptomic, genetic, and biological data. Utilising the multi-omic network inference method KiMONO (16), we incorporated GR-meSNPs (n=88,585), biological variables such as sex, age, BMI, case-control status, and predicted white BCC, GR-meCPGs (n=3,772), and all available transcripts (n=11,994). For network generation, we used the changes in DNAm and gene expression. In the inferred multi-level GR-network, nodes represent genes, SNPs, CpG sites, or biological variables, while connections denote statistically identified effects between them. To ensure the robustness of the network, we excluded node models with low goodness of fit (R2<0.1), absolute effects weaker than beta < 0.04, and stability selection scores less than 100% (k=100 runs). The final GR-network consists of 7,193 nodes connected via 30,332 edges (see Table S6).

We further examined GR-network trios (Fig. 5a), which consist of meQTLs acting as both eQTLs and eQTMs, forming SNP-CpG-transcript trios. By analysing the network relations between transcripts and CpG sites (eQTM) and between transcripts and SNPs (eQTLs), we identified 552 eQTMs and 297 eQTLs mapping back to 7,979 GR-meQTLs (Fig. 5b and Table S7). Additionally, we discovered 7,591 GR-trio SNPs, 334 GR-trio CpGs, and 613 GR-trio genes. The hub in the network represents a single CpG (cg09614808) located in an open sea region of the gene *Amyloid precursor protein* (*APP*) on chromosome 21, connected with 164 genes distributed across all 22 autosomes. The trio genes displayed enrichment in immune-related pathways such as myeloid leukocyte-mediated immunity, cell activation involved in immune response, myeloid leukocyte activation, as well as pathways related to cell activation, cell motility, and locomotion (see Fig. 5c). Moreover, the trio genes exhibited a significant overlap with genes regulated by dexamethasone administration in the mouse brain using our data published in (45) (n=100 genes, Odds Ratio: 1.8, p-value: 1.4x10^-6^, refer to Fig. 5d, Table S8). Importantly, 59% of the overlapping genes were expressed in multiple brain regions, with the prefrontal cortex showing the highest degree of overlap (Fig 5e). These findings underscore the potential importance of these trio genes in the context of the impact of stress in the brain and their potentially critical role in mediating the impact of glucocorticoids on brain function and behaviour.

### Supplementary Tables

**Table S1:** List of 3,280 significant GR-DMPs (Differentially Methylated CpG Sites after dexamethasone treatment compared to baseline) showing CpG site identifiers (CpG), statistical significance (P-value), chi-squared statistics (Chi_sq), fold change (FC), variance (Var), false discovery rate (FDR), chromosome (Chr), genomic position (Pos), relation to CpG island (Relation_to_Island), and UCSC RefGene name (UCSC_RefGene_Name).

**Table S2**: List of 104,828 significant GR-meQTLs. The table includes CpG site identifiers (CpG), single nucleotide polymorphism (SNP) identifiers, beta values (Beta), t-statistics (T-stat), p-values (P-value), false discovery rates (FDR), chromosome (Chr), SNP position (SNP_pos), CpG start position (CpG_start), CpG end position (CpG_end), and the relation of the CpG to a CpG island (Relation_to_Island).

**Table S3**: List of 7,022,984 significant baseline meQTLs. The table includes CpG site identifiers (CpG), single nucleotide polymorphism (SNP) identifiers, beta values (Beta), t-statistics (T-stat), p-values (P-value), false discovery rates (FDR), chromosome (Chr), SNP position (SNP_pos), CpG start position (CpG_start), CpG end position (CpG_end), and the relation of the CpG to a CpG island (Relation_to_Island).

**Table S4**: List of 28,688 significant GR-eQTMs, including CpG site identifiers (CpG), transcript identifiers (Transcript), beta values (Beta), t-statistics (T-stat), p-values (P-value), false discovery rates (FDR), chromosome (Chr), transcript start position (Transcript_start), transcript end position (Transcript_end), gene symbol associated with the transcript (Transcript_Gene_symbol), CpG position (CpG_Pos), and CpG relation to UCSC RefGene (CpG_UCSC_RefGene_Name). These associations represent GR-induced relationships between DNA methylation and gene expression in cis-regulatory regions.

**Table S5:** List of 1,307 significant eQTMs at baseline, including CpG site identifiers (CpG), transcript identifiers (Transcript), beta values (Beta), t-statistics (T-stat), p-values (P-value), false discovery rates (FDR), chromosome (Chr), transcript start position (Transcript_start), transcript end position (Transcript_end), gene symbol associated with the transcript (Transcript_Gene_symbol), CpG position (CpG_Pos), and CpG relation to UCSC RefGene (CpG_UCSC_RefGene_Name).

**Table S6**: Characteristics of the GR network. The network comprises 7,193 nodes and 30,332 edges, constructed using KiMONo. The table includes information about the network nodes, specifically the target node (Target) and predictor node (Predictor), along with statistics such as the mean and standard deviation (SD) of various network properties, including mean RSQ, SD RSQ, mean MSE, and SD MSE. Additionally, information about the predictor and target layers within the network is provided.

**Table S7**: Characteristics of the GR-network trios, which consist of meQTLs acting as both eQTLs and eQTMs. The table includes information about the network nodes, specifically the target node (Target) and predictor node (Predictor), along with statistics such as the mean and standard deviation (SD) of various network properties, including mean RSQ, SD RSQ, mean MSE, and SD MSE. Additionally, information about the predictor and target layers within the network is provided.

**Table S8**: List of trio genes significantly overlapping with genes regulated by dexamethasone administration in the mouse brain. The table includes identifiers for both mouse and human genes, such as Mouse Ensembl ID, Mouse Gene Symbol, Human Ensembl ID, Human Gene Symbol, and information on specific brain regions.

**Table S9**: GR-trio variants, which include 321 LD independent SNPs. This table provides information on the chromosome (CHR), SNP position (Pos), meQTL FDR, total number of SNPs, and clumped SNPs.
